# Impact of Anti-VEGF Treatment for Diabetic Macular Oedema on Progression to Proliferative Diabetic Retinopathy: Data-driven Insights from a Multicentre Study

**DOI:** 10.1101/2023.11.10.23298261

**Authors:** Abraham Olvera-Barrios, Watjana Lilaonitkul, Tjebo FC Heeren, Assaf Rozenberg, Darren Thomas, Alasdair N. Warwick, Taha Somroo, Abdulrahman H. Alsaedi, Roy Schwartz, Usha Chakravarthy, Haralabos Eleftheriadis, Ashish Patwardhan, Faruque Ghanchi, Paul Taylor, Adnan Tufail, Catherine Egan, UK DR EMR Users group

## Abstract

**Background:** To report insights on proliferative-diabetic-retinopathy (PDR) risk modification with repeated anti-vascular endothelial-growth-factor (VEGF) injections for the treatment of diabetic-macular-oedema (DMO) in routine care, and present data-driven PDR screening recommendations for injection clinics.

**Methods:** Multicentre study (27 UK-NHS centres) of patients with non-PDR with and without DMO. Primary outcome was PDR development. Repeated anti-VEGF injections were modelled as time-dependent covariates using Cox regression and weighted cumulative exposure (WCE) adjusting for baseline diabetic retinopathy (DR) grade, age, sex, ethnicity, type of diabetes, and deprivation. A propensity score matched cohort was used to estimate the treatment effect on PDR incidence rates (IR).

**Results:** We included 5716 NPDR eyes (5716 patients, 2858 DMO eyes). The WCE method showed a better model fit. Anti-VEGF injections showed a protective effect on risk of PDR during the most recent 4-weeks from exposure which rapidly decreased. There was a 20% reduction in risk of PDR (p0.006) in treated eyes. Severe-NPDR had a 4.6-fold increase in PDR hazards when compared with mild-NPDR (p<0.001). The annual IR of untreated mild-NPDR cases was 2.3 [95%CI 1.57-3.23] per 100 person-years). In NPDR DMO cases treated with anti-VEGF, similar IR would occur with annual review for mild, 6-monthly for moderate, and 3-monthly for severe-NPDR.

**Conclusion:** The WCE method is a better modelling strategy than traditional Cox models for repeated exposures in ophthalmology. Injections are protective against PDR predominantly within the most recent 4 weeks. Based on observed data, we suggest follow-up recommendations for PDR detection according to retinopathy grade at first injection.

**Précis:** This study describes the impact on PDR risk of anti-VEGF injections for DMO in routine care and data-driven reassessment recommendations of the peripheral retina for people in long term injection clinics.

**Key messages:** *What is already known on this topic:*

– Clinical trials have shown that intravitreal anti-vascular endothelial growth factor (VEGF) injections reduce the incidence rate of proliferative diabetic retinopathy (PDR).
– Repeated intravitreal anti-VEGF injections are the mainstay of treatment for diabetic macular oedema (DMO), however, there is little evidence on how these exposures impact on the risk of PDR in clinical practice.

*What this study adds:*

– The impact of anti-VEGF on PDR risk varies based on the timing of exposure and the effect is not permanent.
– Despite repeated treatments with anti-VEGF injections, patients with DMO may still progress to PDR.

*How this study might affect research, practice, or policy:*

– Our work underscores the significance of taking into account repeated treatments at varying time intervals in ophthalmology, highlighting the utility of the weighted cumulative exposure method.
– Implementing adequate modelling strategies to address the complexities of exposures in clinical settings can improve predictions and patient outcomes.
– We provide PDR screening recommendations for DMO patients undergoing anti-VEGF treatments in injection clinics. Implementation would improve the safety and efficiency of treatment pathways.

## Introduction

Proliferative diabetic retinopathy (PDR) and diabetic macular oedema (DMO) are among the leading causes of incident sight impairment and blindness in the working age population.[1,2]

Treatment of DMO and PDR has evolved over the last few decades after pivotal laser treatment trials.[3,4] Anti-vascular endothelial growth factor (VEGF) therapy has become the standard of care for managing DMO.[5] Despite rigorous and intensive intravitreal anti-VEGF DMO treatment protocols in clinical trials, progression to PDR occurs in treated eyes, though at lower rates.[4–6] Moreover, preventive treatment with anti-VEGF in patients with non-PDR and no DMO, shows that DMO and/or PDR incidence risk is not eliminated.[4,5,7,8] PDR risk over time was estimated by the Early Treatment Diabetic Retinopathy study (ETDRS), defining follow-up intervals (pre-dating anti-VEGF therapies).[9,10] Little is known about how treatment regimens impact PDR risk outside clinical trials, whether the effect of repeated anti-VEGF injections accumulates or persists after treatment cessation, and whether the same PDR surveillance recommendations apply for treated eyes. Accounting for the exposure to anti-VEGF injections is important in the context of DR. Anti-VEGF injections reduce DR progression rates and improve DR severity score (DRSS).[4,7,8,11] Estimating the effect of intravitreal anti-VEGF for DMO treatment on PDR development with real-world data is challenging due to confounding, variability in treatment regimens, uncertainty about cumulative effect, and the uncertain relevance of injections given at different time intervals. An appropriate representation of time-dependent exposures is necessary to avoid underestimating the effect.[12] Novel methods model the history of drug exposure, such as the weighted cumulative exposure (WCE), flexibly representing the exposure’s past effect on current risk through recency-weighted exposures.

The present study addresses a significant gap in our knowledge by utilising real-world, rather than modelled, data to examine the impact of anti-VEGF exposures on progression to PDR in individuals where the indication for anti-VEGF injections was DMO. Additionally, we provide data-driven PDR screening interval recommendations for patients receiving courses of intravitreal injections, which may last for years, and may occur in treatment pathways where the peripheral retina is not examined at every clinic visit.

## Methods

### Study design and setting

This was a retrospective multicentre study. The UK DR EMR User Group gathers large-scale routine clinical data to improve the prevention, diagnosis and treatment of DR. Twenty-seven UK centres using the same EMR system (Medisoft, Medisoft-Ltd, Leeds UK), with mandated structured DR feature grading and time stamped interventions (e.g. intravitreal injections, laser procedures, and ophthalmic surgeries), contributed data. An algorithm generates DR grades according to ETDRS,[10] International,[13] and English National Screening Committee (NSC-UK)[14] DR classification systems. Each site is the only English National Health Service provider of DR care for the local population and very few patients switch between providers or access private care. All patients were new referrals from the English diabetic eye screening program (DESP), a nationwide systematic program maintained by rigorous quality assurance measures.

### Data extraction

Anonymised data were remotely extracted through the EMR’s DR module from the time of their first DR structured assessment entry onto the EMR, to the date of their last clinical entry before the data extraction on 31-December-2018.[15] Demographic data were extracted from the hospital’s patient administration system to the EMR. The cohort went through a staged exclusion process (supplementary-figure 1) to identify anti-VEGF treated and treatment-naive eyes with similar characteristics. Eyes with PDR at baseline or at time of first injection were excluded. Treated patients who did not complete a loading dose of at least 3 anti-VEGF injections and patients with indications for anti-VEGF treatment other than DMO were excluded.

The recording of clinical variables has been described elsewhere,[16] and included age, sex, type of diabetes (categorised as type 1, type 2 and other), ethnicity (categorised as White, Black, Any other Asian, Other and not stated) and deprivation, measured as the index of multiple deprivation (IMD), the official measure of relative deprivation in England.[17] Composite ETDRS scores were automatically generated in the EMR. For analysis, all eyes were graded at clinic visits as mild-NPDR (ETDRS levels 20-35), moderate-NPDR (ETDRS level 43), severe-NPDR (ETDRS levels 47-53) and PDR (ETDRS levels 61-81).

### Statistical analysis

The main outcome measure was PDR development, defined as retinal or optic nerve new vessels (NV), NV of the iris or angle, neovascular glaucoma, tractional retinal detachment, vitreous haemorrhage, preretinal haemorrhage or record of pan-retinal photocoagulation or vitrectomy for PDR-related reasons. One eye per patient was included for analysis in the treated cohort as follows: 1) the first treated eye; 2) if the first treatment was bilateral, the eye with worse DR grade was selected; and 3) if the first treatment was bilateral and the DR grade was the same bilaterally, a random selection was carried out. In treatment-naive eyes, the eye with worse DR grade was included, and if the DR grade was the same bilaterally, a random selection was carried out. Eyes were censored on change in their treatment modality (other than anti-VEGF), intraocular surgery, or their last DR assessment/follow-up. Survival curves were generated using the Turnbull estimator.

### Effect of past anti-VEGF exposures on risk of PDR

To examine the impact on model fit when accounting for repeated exposures in treated eyes, we implemented 3 different modelling strategies with different degrees of complexity using Cox regression and the novel WCE method.[18] The proportionality assumption was tested graphically by inspection of Schoenfeld residuals.[19] All models adjusted for baseline DR severity, age, sex, ethnicity, type of diabetes, and IMD. We compared goodness of fit using Akaike information criterion (AIC) while accounting for differences in degrees of freedom.[18,20] Lower AIC values indicate a better fit. Differences in AIC above 10 units are considered significant.[20] Briefly, a simpler model with higher AIC (worse fit) would likely suffer from under-fit bias when compared to a more complex model with a lower AIC (better fit).[21]

A traditional Cox_model_ adjusted for all time-fixed covariates and ignored anti-VEGF exposures. Cox_model_ allowed comparisons with models adjusting for anti-VEGF injections. Using an extension of the Cox model (Cox_tdc_), anti-VEGF treatments were modelled as a time-dependent unweighted cumulative sum of exposures.[19,22] In pharmacoepidemiology, current risk may be affected by a recent increase in exposures.[12] Due to uncertainty about how past anti-VEGF injections impact PDR risk, we used weight functions fitted to the data using flexible restricted cubic regression splines modelled in Cox regression, avoiding a priori assumptions about the specific shape of the weight function following the methods from Sylvestre et al.[18] (WCE_model_). We considered 5 different windows of etiologically relevant exposure. For each time window, alternative models, with 1 to 3 interior knots uniformly placed across the time window were compared. The length of time window before index date and number of internal knots were chosen from the model with best fit assessed by AIC (where minimum AIC means a better fit).[18] The linear combination of estimates from the weight function was used to calculate a WCE score for each individual. The WCE_model_ controlled for time-fixed covariates and for each patient’s WCE score.

### Propensity score matching

We used propensity score matching (PSM) in a treatment-naive DR cohort without DMO adjusting for age, sex, baseline DR severity, and IMD. A 1:1 nearest neighbour PSM without replacement with a propensity score estimated using logistic regression of the treatment on the covariates resulted in adequate balance (supplementary-figure 2). The effect of anti-VEGF injections on the treated was estimated with a Cox model with matching weights applied. Robust standard errors were used for 95% CI and p-value calculations.

### Follow-up recommendations and anti-VEGF impact on risk of PDR

We calculated PDR cumulative incidence rates (IR) per 100 person-time (where time is equal to the screening interval assessed). We computed IRs at 3, 6, 12, and 24 months for each DR severity level. To ensure equitable delivery of eye care, IRs were compared with IRs observed in the treatment-naive cohort. Following UK-NSC[23] and American Diabetes Association[24] DR screening recommendations, and considering follow-up in a tertiary, rather than a community setting, we set the PDR IR corresponding to the annual IR of treatment-naive eyes with mild-NPDR as reference. IR, rate ratio, and absolute difference in IR were calculated to provide further insights on the impact of anti-VEGF treatment on PDR incidence.

As sensitivity analysis, IR, rate ratios, and absolute differences in IR were calculated in an unmatched treatment-naive cohort.

All analyses were performed using R version 4.0.3. The survival,[25] and WCE[18] packages were used for survival analyses. The MatchIt[26] package was used for PSM.

## Results

A total of 5716 eyes (2858 treated vs 2858 treatment-naive eyes) from 5716 patients (61% Male, 3489/5716) with NPDR were included (supplementary-figure 1). Table 1 shows cohort characteristics; 54% (3107/5716) of patients self-described as white, and more than a third (32%, 1830/5716) were in the most deprived quintile of deprivation (first quintile). There were 209 incident PDR cases over a median (IQR) follow-up of 1.2 (0.6-2.4) years in the treated cohort (4.45 [95%CI 3.89-5.09] PDR cases per 100 person-years). The median (IQR) number of intravitreal injections was 11 (7-19); 11 (6-18) for mild-NPDR eyes, 12 (7-18) for moderate-NPDR, and 12 (7-23) for severe-NPDR. The median number of injections by baseline DR severity per year of follow-up is shown in supplementary-table 1. Most patients received ranibizumab (67%, 1921/2858) at baseline, followed by aflibercept (31%, 889/2858), and bevacizumab (2%, 48/2858). In the treatment-naive cohort, there were 537 incident PDR cases over a median (IQR) follow-up of 2.44 (1.19-4.45) years (6.27 [95%CI 5.77-6.81] PDR cases per 100 person-years).

**Table 1.** Cohort characteristics.

| Characteristic | Group |  |  |
| --- | --- | --- | --- |
|  | Overall, N = | Treated, N = | Treatment-naïve, N = |
|  | 5,716 | 2,858 | 2,858 |
| **Age** | 64 (57, 72) | 64 (57, 72) | 64 (57, 72) |
| **Sex** |  |  |  |
| Female | 2,227 (39%) | 1,115 (39%) | 1,112 (39%) |
| Male | 3,489 (61%) | 1,743 (61%) | 1,746 (61%) |
| **Baseline DR grade** |  |  |  |
| Mild NPDR | 926 (16%) | 463 (16%) | 463 (16%) |
| Moderate NPDR | 3,872 (68%) | 1,935 (68%) | 1,937 (68%) |
| Severe NPDR | 918 (16%) | 460 (16%) | 458 (16%) |
| **Ethnicity** |  |  |  |
| White | 3,107 (54%) | 1,660 (58%) | 1,447 (51%) |
| South Asian | 654 (11%) | 313 (11%) | 341 (12%) |
| Black | 373 (6.5%) | 209 (7.3%) | 164 (5.7%) |
| Any other Asian | 125 (2.2%) | 61 (2.1%) | 64 (2.2%) |
| Other | 122 (2.1%) | 71 (2.5%) | 51 (1.8%) |
| Not stated | 1,335 (23%) | 544 (19%) | 791 (28%) |
| **Type of diabetes** |  |  |  |
| Type 2 | 4,057 (71%) | 2,129 (74%) | 1,928 (67%) |
| Type 1 | 436 (7.6%) | 183 (6.4%) | 253 (8.9%) |
| Other | 169 (3.0%) | 66 (2.3%) | 103 (3.6%) |
| Unknown | 1,054 (18%) | 480 (17%) | 574 (20%) |
| **IMD (quintiles)** |  |  |  |
| 1 | 1,830 (32%) | 915 (32%) | 915 (32%) |
| 2 | 1,381 (24%) | 688 (24%) | 693 (24%) |
| 3 | 956 (17%) | 479 (17%) | 477 (17%) |
| 4 | 764 (13%) | 383 (13%) | 381 (13%) |
| 5 | 785 (14%) | 393 (14%) | 392 (14%) |

Median (IQR) for continuous variables.
Count (column %) for categorical variables.

### Effect of past anti-VEGF exposures

The WCE_model_ with a regression spline function based on three equally spaced knots over a six-month time window exposure showed the best fit to the data when compared with Cox_model_ and Cox_tdc_ (AIC=2837.6, supplementary-tables 2, 3). Supplementary-figure 3 shows the estimated weight function for the WCE_model_, which illustrates the relative strength of anti-VEGF impact on PDR risk. Current and most recent doses showed the highest protective effect (negative weights). This effect rapidly decreases with increasing time after exposure, reaching 0 at approximately week 4. More remote doses did not seem to have an impact on current risk of PDR.

We report mutually adjusted hazard ratios (HR) from the WCE_model_ (best fitting model, table 2). The poorest fit to the data was seen in Cox_model_ which ignored the exposure of intravitreal injections (AIC difference of 94 vs WCE_model_). Mutually adjusted HR from Cox_model_ and Cox_tdc_ are available in supplementary-table 4.

**Table 2.** Mutually adjusted hazard ratios allowing for baseline diabetic retinopathy grade, age, sex, type of diabetes, ethnicity, and index of multiple deprivation. Anti-VEGF exposures are modelled as cumulative exposures weighted by recency.

| Characteristic | HR (95% CI)* | p-value |
| --- | --- | --- |
| **Baseline DR grade** |  |  |
| Mild NPDR | 1.00 |  |
| Moderate NPDR | 1.99 (1.13, 3.51) | **0.015** |
| Severe NPDR | 4.63 (2.55, 8.41) | **2.8e-07** |
| Age (per 5-year rise) | 0.91 (0.85, 0.96) | **0.002** |
| **Sex** |  |  |
| Female | 1.00 |  |
| Male | 1.36 (1.00, 1.84) | **0.047** |
| **Type of diabetes** |  |  |
| Type 2 | 1.00 |  |
| Type 1 | 2.08 (1.35, 3.21) | **7.3e-04** |
| Other | 1.24 (0.50, 3.10) | 0.639 |
| Unknown | 0.89 (0.60, 1.32) | 0.552 |
| **Ethnicity** |  |  |
| White | 1.00 |  |
| South Asian | 0.91 (0.58, 1.45) | 0.698 |
| Black | 0.44 (0.20, 0.96) | **0.036** |
| Any Other Asian | 0.80 (0.29, 2.21) | 0.657 |
| Other | 0.34 (0.08, 1.42) | 0.132 |
| Not Stated | 1.02 (0.70, 1.48) | 0.917 |
| **IMD quintiles** |  |  |
| 1 (most deprived) | 1.00 |  |
| 2 | 1.03 (0.72, 1.49) | 0.867 |
| 3 | 1.03 (0.68, 1.56) | 0.903 |
| 4 | 0.87 (0.56, 1.36) | 0.542 |
| 5 (least deprived) | 0.62 (0.37, 1.04) | 0.064 |

\*HR; hazard ratio, CI; confidence interval

### Progression to proliferative diabetic retinopathy

Figure 1 shows PDR development probabilities stratified by baseline DR severity. At year 4, the probability of PDR was 24.4% in the treatment-naive eyes and 14.8% in the treated eyes. The effect of anti-VEGF treatment estimated with a Cox PH model with PSM weights applied showed a 20% reduction in hazards of PDR (HR 0.80, 95%CI 0.68-0.94, p0.006). Supplementary-table 5 shows the differences in IRs from observed data. On average, there were almost 2 fewer incident PDR cases per 100 persons-year in treated eyes. The difference was most notable among people with severe-NPDR (rate ratio 0.65, 95%CI 0.45-0.93). Differences were not significant between mild-NPDR eyes.

**Figure 1.**
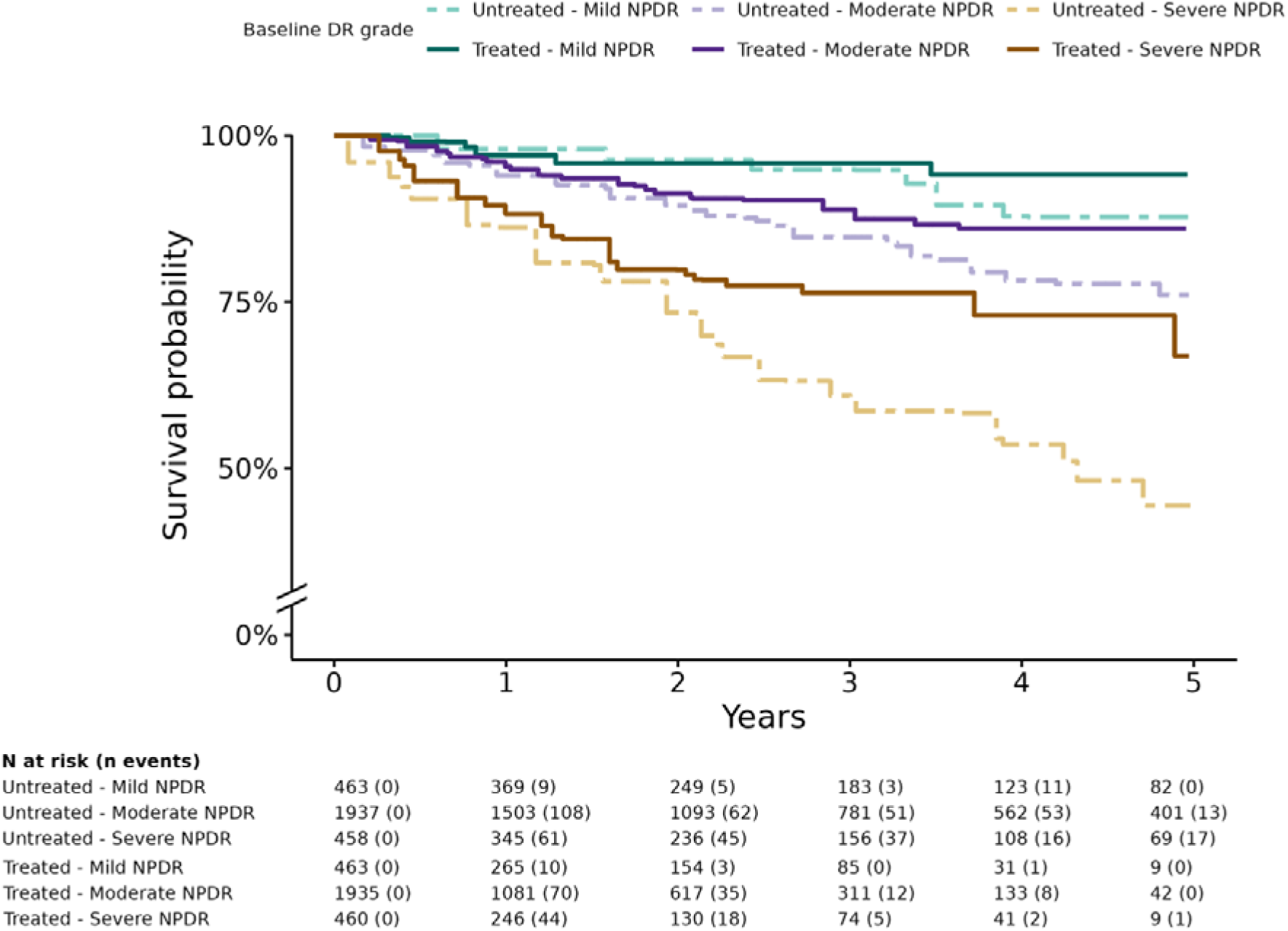
Survival curves for progression to proliferative diabetic retinopathy (PDR) stratified by baseline diabetic retinopathy (DR) severity.

Sensitivity analyses comparing IR in unmatched treatment-naive eyes (53376 treatment-naive DR eyes vs 2858 DMO anti-VEGF treated eyes) showed consistent differences in IR and rate ratios in eyes with mild and moderate-NPDR (supplementary-table 6). Differences were not significant overall, possibly owing to observed imbalances in DR severity and age (older population with higher proportion of mild-NPDR cases in treatment-naive eyes, supplementary-figure 2).

### Data-driven follow-up recommendations for PDR screening

The cumulative PDR IR in treatment-naive patients with mild-NPDR was 2.26 (95%CI 1.57-3.23) per 100 person-years. This would mean that, on average, 2 new PDR cases per 100 persons occur over a one-year period. Allowing for the occurrence of 2 new PDR cases per period, yields a recommended PDR interval screening of 12 months for mild-NPDR (IR 1.81, 95%CI 1.03-3.09), 6 months for moderate-NPDR (IR 1.97, 95%CI 1.65-2.35), and 3 months for severe-NPDR cases (IR 2.35, 95%CI 1.85-2.97) with DMO treated with anti-VEGF. Figure 2 shows IRs by pre-specified screening interval.

**Figure 2.**
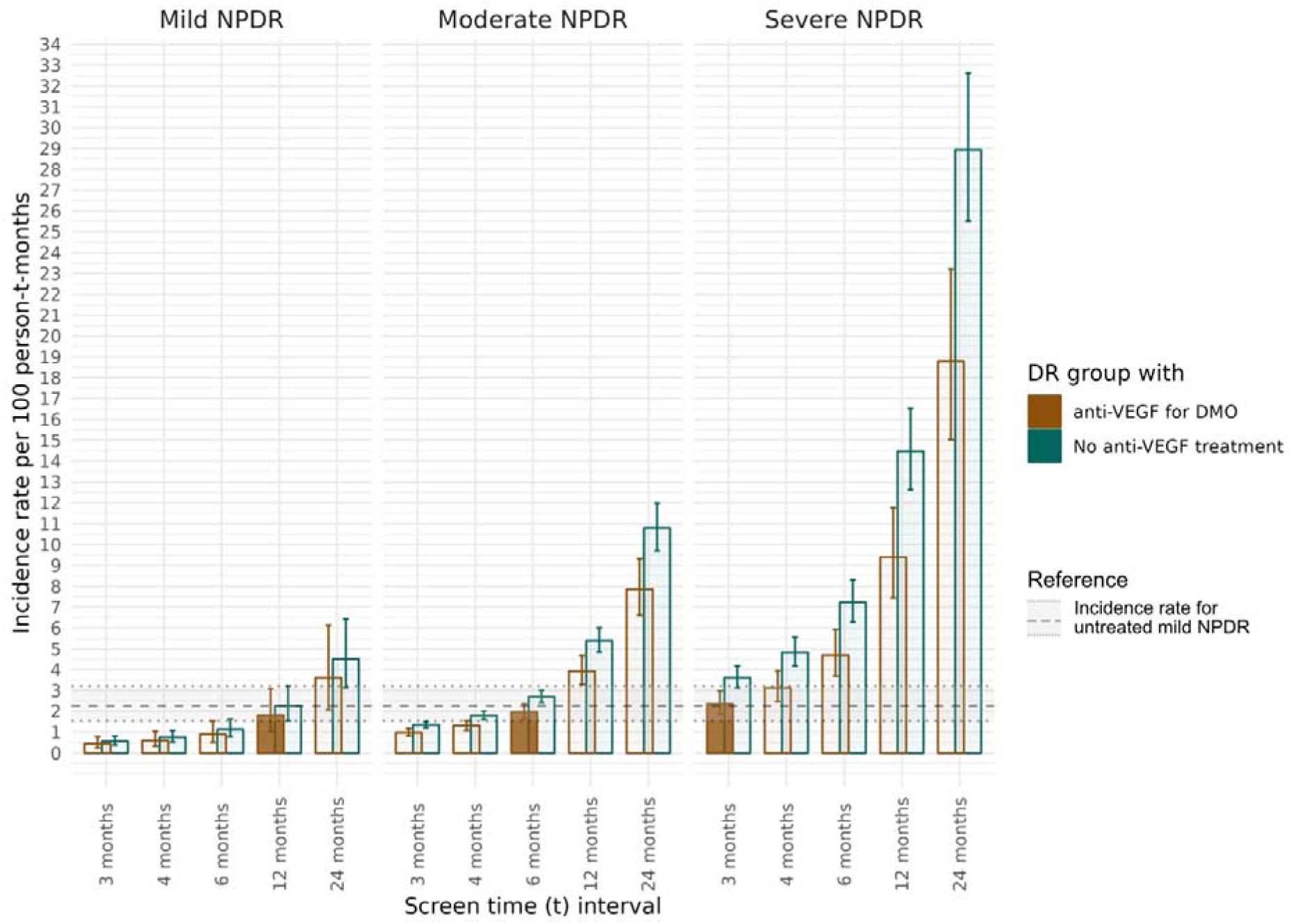
Cumulative incidence rates of proliferative diabetic retinopathy at different time periods to drive screening recommendations. Time (*t)* tests the different screening periods for each diabetic retinopathy severity levels which vary along the x axis.

### Associations with progression to proliferative diabetic retinopathy

Eyes with severe-NPDR at baseline showed a more than 4-fold increase in PDR hazards (HR 4.63, 95%CI 2.55-8.41, p2.8×10^-7^) when compared with mild-NPDR patients (table 2). Every 5-year rise in age showed a 9% reduction in PDR hazards (95%CI 0.85-0.96; p6.0×10^-4^). PDR hazards in patients with Type 1 diabetes were 2.08-fold higher when compared with type 2 diabetes patients (p7.3×10^-4^). Compared with females, males showed a 36% increase in PDR hazards (p0.047). Black patients showed a 56% reduction in PDR hazards when compared with white participants.

In a DR feature-based sub-analysis of treated eyes with severe-NPDR, only intraretinal-microvascular-abnormalities (IRMA) showed a significant association with PDR (HR 2.55, 95%CI 1.12-5.79, *p*=0.022) when compared with dot-blot haemorrhages in 4 quadrants (supplementary-results, supplementary-table 7).

## Discussion

This study provides a comprehensive analysis of PDR development in DMO eyes undergoing anti-VEGF treatment in routine clinical practice. Our findings support the use of WCE to model the complex time-dependent exposures to anti-VEGF injections, preventing underfitting in real-world data. Our research sheds light on the duration of the protective effect of anti-VEGF injections particularly during the most recent 4 weeks from treatment (supplementary-figure 3). Despite frequent intravitreal injections, DMO eyes treated with anti-VEGF still develop PDR, but showed a 20% reduction in the risk of PDR when compared with treatment-naive eyes (figure 1). Based on our findings, we propose evidence-based intervals for peripheral retinal examination for new onset neovascularisation based on the baseline DR grade: likely to be important for “injection-only” clinical pathways.

### The effect of intravitreal anti-VEGF injections on PDR progression

Incorporating intravitreal anti-VEGF exposures as weighted time-dependent covariates yielded a far better fit to our data (supplementary-table 3), providing evidence that PDR risk is modified by anti-VEGF exposures and varied with treatment recency. The RISE and RIDE extension study[6] demonstrated that patients with DRSS stability/improvement received more injections compared to those with worsened scores (p<0.001). Furthermore, greater instability occurred in more severe baseline cases (p<0.001).[6] The PANORAMA study suggested a dose-dependent effect on DRSS improvement,[8] supporting the importance of past anti-VEGF exposures and baseline DR severity. The protective anti-VEGF effect declines sharply, becoming insignificant at approximately 4 weeks after exposure. Reduction in treatment frequency to less-than-monthly injections, or treatment discontinuation, translates to an increase in PDR risk compared with eyes treated with monthly injections which is even greater in high-risk groups.

### Progression to proliferative diabetic retinopathy

Nguyen et al.[27] simulated treating severe-NPDR with rigorous early anti-VEGF injections versus delaying treatment until PDR development using extrapolated 1-year data from PANORAMA[8] and RISE/RIDE[28] trials. Based on real-world data from over 77000 patients, a 2 million NPDR patient cohort was simulated. Early treatment showed a 51.7% relative risk reduction in PDR development over 5 years and a 19.4% absolute risk reduction. The authors highlight the lack of real-world evidence of anti-VEGF treatment on risk of PDR. However, as only 1-year RCT data is extrapolated in a Monte Carlo model, no differences in the timing and frequency of the treatments were taken into account, and results are likely to differ in real-world settings where intensity and timing of treatment vary. Our study addresses this by showing a 20% average reduction effect in PDR risk in the treated population (p0.006), and by providing real-world pharmacoepidemiologic evidence of the importance of past anti-VEGF treatments (supplementary-figure 3).

The PANORAMA study[8] studied the effect of longer anti-VEGF treatment intervals for patients with no DMO and showed a PDR cumulative probability within 2 years of 13.5% for treated eyes and 33.2% for sham eyes. In the RISE/RIDE studies,[4] 8% (21/257) of cases in the sham group progressed to PDR at 2 years follow-up from NPDR at baseline, compared with 2% (10/502) of those treated with ranibizumab. Total progression of retinopathy in RISE/RIDE by all measures was 30% (78/257) in the sham group and 10% (51/502) in the treated groups. The DRCRnet protocol W[7] explored the potential use of anti-VEGF injections at earlier stages of retinopathy to prevent DR progression in eyes without DMO, finding 2-year PDR cumulative probability of 13.5% in the aflibercept treated group. The PDR cumulative probability at year-2 in our study was 12.2% in the treatment-naive vs 9.7% in the treated cohort. Possible reasons for the lower PDR cumulative incidence in our cohort at year-2 are the variation in appointment intervals in routine care and differences in baseline characteristics. Unlike clinical trials, we provide cumulative PDR risk estimates for up to 5 years of follow-up.

We recommend review under mydriasis for NPDR eyes with DMO undergoing anti-VEGF treatment every 12-months for mild-NPDR, 6-monthly for moderate-NPDR, and 3-monthly for severe-NPDR. Although in the long term, novel grading systems incorporating widefield optical coherence tomography-angiography, may be resilient to the effect of anti-VEGF in their predictive power and may eventually supplant the ETDRS grading system, it will take many years to curate datasets to validate such systems. Currently, in the absence of novel grading systems, a pragmatic guide to surveillance is needed given the prevalence of injection treatments for DMO.[29]

### Proliferative diabetic retinopathy associations

Baseline DR severity is one of the most relevant biomarkers for predicting patient outcomes, even with frequent anti-VEGF injections. Baseline DR severity grade was associated with 1.22 and 4.63-fold rises in PDR hazards for moderate and severe-NPDR when compared with mild-NPDR, respectively. IRMA showed a 2.55-fold risk of progressing to PDR when compared with dot-blot-haemorrhages in 4 quadrants, further highlighting the relevance of clinical DR features.[9,10,16]

Younger patients were at increased risk for developing PDR, confirming the findings of other studies.[30,31] Males showed a 36% increase in PDR hazards when compared with females. Previous studies on DR and DMO have not shown significant sex differences. Sex is a known confounder in diseases involving the microvasculature,[16,32] and together with evidence of a reduced likelihood of achieving metabolic therapeutic goals,[30] are possible reasons for the observed differences. Black patients have been reported to be more likely to develop DR than white patients, more likely to present with sight-threatening DR,[33,34] and less likely to attend for diabetic eye screening.[35] Interestingly, white patients showed worse survival compared with black patients. It is worth noting that 19% (544/2858) of patients had missing ethnicity data, underscoring the need for caution in interpretation of ethnic associations. Type 1 diabetes and worse DR severity at baseline were the strongest predictors of PDR development.

### Strengths and limitations

This study has several strengths. Firstly, we examined a large cohort with structured data collection in EMR including retinopathy grade, retinal features, time-stamped diagnoses, clinical examinations, and treatments/procedures. Secondly, we have analysed the relative importance of intravitreal injections given in routine clinical care settings, rather than modelled data, on PDR development by modelling injections as weighted exposures in Cox regression using the novel WCE method. Thirdly, we have used PSM to estimate the average effect of anti-VEGF treatment on the risk of PDR.

The main limitations of our study are as follows. Our data are dependent on the quality of EMR examination and recordings. Large epidemiologic or clinical studies put constraints either on participants and on researcher behaviour and are increasingly more difficult and costly to perform; the results of real-world practice provide equally important, but different, information, where the scale of the dataset compensates in part for the potential for inaccuracies in the clinical record. A second limitation of our study is that data on confounders such as level of hyperglycaemia, hypertension, and other systemic comorbidities was not available. As with any observational study, results can be influenced by residual unaccounted confounders that can only be resolved by a randomised controlled trial of differing intensity and duration of treatment regimens.

## Conclusion

Our results support the use of WCE in modelling the effects of anti-VEGF injections in routine clinical practice and demonstrate that eyes with repeated anti-VEGF treatment for DMO still progress to PDR. Our WCE model suggests that intravitreal anti-VEGF therapy is associated with a reduction in risk of PDR which lasts for 4 weeks after each injection. Baseline DR grade, clinical retinopathy features (IRMA), age, sex, type of diabetes, and ethnicity were key prognostic factors. Our findings highlight the risk of PDR progression after intravitreal anti-VEGF therapy has been initiated, or stopped, and are, together with our pragmatic screening interval recommendations, of clinical significance for the management and monitoring of DMO patients.

## Contributors

AO-B, WL, PT, CE, and AT designed the study. CE, AT, PT, UC, HE, AP, FG, participated in data acquisition. AO-B, DT, and TH curated the data. AO-B, WL, TH, and DT undertook statistical analyses. AO-B, WL, TH, DT, and PT had direct access and verified the underlying data. AO-B, and WL wrote the first version of the manuscript. All authors interpreted results, critically reviewed, and edited the manuscript. AO-B, PT, and CE had the final responsibility for the decision to submit for publication and are guarantors of the work.

## Ethics approval

The study was conducted in accordance with the Declaration of Helsinki and the United Kingdom Data Protection Act. The lead clinician and Caldicott Guardian responsible for protecting the confidentiality of patient information, at each centre, gave written approval for extraction of anonymised data. The study protocol was approved by the head of research governance at Moorfields Eye Hospital NHS Foundation Trust, the lead clinical centre.

## Data availability statement

Owing to the comprehensive multicentre electronic medical record data collection, the tables required to reproduce the aforementioned findings cannot be shared at this time due to ethical considerations.

## Meeting presentation

Presented at the Association for Research and Vision in Ophthalmology (ARVO) annual meeting, 2021.

## References

1 Sun H, Saeedi P, Karuranga S, et al. IDF Diabetes Atlas: Global, regional and country-level diabetes prevalence estimates for 2021 and projections for 2045. Diabetes Res Clin Pract 2022;183:109119.

2 Olvera-Barrios A, Mishra AV, Schwartz R, et al. Formal registration of visual impairment in people with diabetic retinopathy significantly underestimates the scale of the problem: a retrospective cohort study at a tertiary care eye hospital service in the UK. Br J Ophthalmol Published Online First: 14 October 2022. doi:10.1136/bjo-2022-321910

3 Bressler SB, Liu D, Glassman AR, et al. Change in Diabetic Retinopathy Through 2 Years: Secondary Analysis of a Randomized Clinical Trial Comparing Aflibercept, Bevacizumab, and Ranibizumab. JAMA Ophthalmol 2017;135:558–68.

4 Ip MS, Domalpally A, Sun JK, et al. Long-term effects of therapy with ranibizumab on diabetic retinopathy severity and baseline risk factors for worsening retinopathy. Ophthalmology 2015;122:367–74.

5 Mitchell P, McAllister I, Larsen M, et al. Evaluating the Impact of Intravitreal Aflibercept on Diabetic Retinopathy Progression in the VIVID-DME and VISTA-DME Studies. Ophthalmol Retina 2018;2:988–96.

6 Goldberg RA, Hill L, Davis T, et al. Effect of less aggressive treatment on diabetic retinopathy severity scale scores: analyses of the RIDE and RISE open-label extension. BMJ Open Ophthalmology 2022;7:e001007.

7 Maturi RK, Glassman AR, Josic K, et al. Four-Year Visual Outcomes in the Protocol W Randomized Trial of Intravitreous Aflibercept for Prevention of Vision-Threatening Complications of Diabetic Retinopathy. JAMA 2023;329:376–85.

8 Brown DM, Wykoff CC, Boyer D, et al. Evaluation of Intravitreal Aflibercept for the Treatment of Severe Nonproliferative Diabetic Retinopathy: Results From the PANORAMA Randomized Clinical Trial. JAMA Ophthalmol 2021;139:946–55.

9 Fundus photographic risk factors for progression of diabetic retinopathy. Ophthalmology 1991;98:823–33.

10 Grading diabetic retinopathy from stereoscopic color fundus photographs--an extension of the modified Airlie House classification. ETDRS report number 10. Early Treatment Diabetic Retinopathy Study Research Group. Ophthalmology 1991;98:786–806.

11 Chiang A, Garg SJ, Klufas MA, et al. Ultra-Response to Ranibizumab: Improvement by 4 or More Steps in Diabetic Retinopathy Severity in Diabetic Retinopathy Clinical Research Network Protocol S. Ophthalmol Retina 2021;5:251–60.

12 Abrahamowicz M, Bartlett G, Tamblyn R, et al. Modeling cumulative dose and exposure duration provided insights regarding the associations between benzodiazepines and injuries. J Clin Epidemiol 2006;59:393–403.

13 Wilkinson CP, Ferris FL 3rd, Klein RE, et al. Proposed international clinical diabetic retinopathy and diabetic macular edema disease severity scales. Ophthalmology 2003;110:1677–82.

14 Harding S, Greenwood R, Aldington S, et al. Grading and disease management in national screening for diabetic retinopathy in England and Wales. Diabet Med 2003;20:965–71.

15 Egan C, Zhu H, Lee A, et al. The United Kingdom Diabetic Retinopathy Electronic Medical Record Users Group, Report 1: baseline characteristics and visual acuity outcomes in eyes treated with intravitreal injections of ranibizumab for diabetic macular oedema. Br J Ophthalmol 2017;101:75–80.

16 Lee CS, Lee AY, Baughman D, et al. The United Kingdom Diabetic Retinopathy Electronic Medical Record Users Group: Report 3: Baseline Retinopathy and Clinical Features Predict Progression of Diabetic Retinopathy. Am J Ophthalmol 2017;180:64– 71.

17 Ministry of Housing, Communities, Local Government. English indices of deprivation 2015. 2015. https://www.gov.uk/government/statistics/english-indices-of-deprivation-2015 (accessed 4 Jul 2023).

18 Sylvestre M-P, Abrahamowicz M. Flexible modeling of the cumulative effects of time-dependent exposures on the hazard. Stat Med 2009;28:3437–53.

19 Tm T, Grambsch PM. Modeling survival data: extending the Cox model. 2000.

20 Quantin C, Abrahamowicz M, Moreau T, et al. Variation over time of the effects of prognostic factors in a population-based study of colon cancer: comparison of statistical models. Am J Epidemiol 1999;150:1188–200.

21 Abrahamowicz M, Ciampi A. Information theoretic criteria in non-parametric density estimation: Bias and variance in the infinite dimensional case. Comput Stat Data Anal 1991;12:239–47.

22 Harrell FE, Jr. Regression Modeling Strategies: With Applications to Linear Models, Logistic and Ordinal Regression, and Survival Analysis. Springer 2015.

23 Diabetic retinopathy. https://view-health-screening-recommendations.service.gov.uk/diabetic-retinopathy/#:∼:text=In%202016%20the%20UK%20NSC,2%20years%20rather%20than%20annually (accessed 20 Jul 2023).

24 Solomon SD, Chew E, Duh EJ, et al. Diabetic Retinopathy: A Position Statement by the American Diabetes Association. Diabetes Care 2017;40:412–8.

25 Therneau T, Lumley T. R survival package. R Core Team Published Online First: 2013. https://rweb.webapps.cla.umn.edu/R/library/survival/doc/survival.pdf

26 Ho DE, Imai K, King G, et al. MatchIt: Nonparametric Preprocessing for Parametric Causal Inference. J Stat Softw 2011;42. doi:10.18637/jss.v042.i08

27 Nguyen QD, Moshfeghi AA, Lim JI, et al. Simulation of long-term impact of intravitreal anti-VEGF therapy on patients with severe non-proliferative diabetic retinopathy. BMJ Open Ophthalmol 2023;8. doi:10.1136/bmjophth-2022-001190

28 Wykoff CC, Eichenbaum DA, Roth DB, et al. Ranibizumab Induces Regression of Diabetic Retinopathy in Most Patients at High Risk of Progression to Proliferative Diabetic Retinopathy. Ophthalmol Retina 2018;2:997–1009.

29 Lundeen EA, Andes LJ, Rein DB, et al. Trends in Prevalence and Treatment of Diabetic Macular Edema and Vision-Threatening Diabetic Retinopathy Among Medicare Part B Fee-for-Service Beneficiaries. JAMA Ophthalmol 2022;140:345–53.

30 Wang L, Li X, Wang Z, et al. Trends in Prevalence of Diabetes and Control of Risk Factors in Diabetes Among US Adults, 1999-2018. JAMA 2021;326:1–13.

31 Hainsworth DP, Bebu I, Aiello LP, et al. Risk Factors for Retinopathy in Type 1 Diabetes: The DCCT/EDIC Study. Diabetes Care 2019;42:875–82.

32 Stratton IM, Kohner EM, Aldington SJ, et al. UKPDS 50: risk factors for incidence and progression of retinopathy in Type II diabetes over 6 years from diagnosis. Diabetologia 2001;44:156–63.

33 Sivaprasad S, Gupta B, Gulliford MC, et al. Ethnic variation in the prevalence of visual impairment in people attending diabetic retinopathy screening in the United Kingdom (DRIVE UK). PLoS One 2012;7:e39608.

34 Olvera-Barrios A, Owen CG, Anderson J, et al. Ethnic Disparities in Progression Rates for Sight-Threatening Diabetic Retinopathy in Diabetic Eye Screening: A Population-Based Retrospective Cohort Study. 2023. doi:10.2139/ssrn.4336087

35 Fathy C, Patel S, Sternberg P Jr, et al. Disparities in Adherence to Screening Guidelines for Diabetic Retinopathy in the United States: A Comprehensive Review and Guide for Future Directions. Semin Ophthalmol 2016;31:364–77.

