## supplements for "Impact of Anti-VEGF Treatment for Diabetic Macular Oedema on Progression to Proliferative Diabetic Retinopathy: Data-driven Insights from a Multicentre Study"

### Supplementary Material

##### Supplementary results

###### Progression to proliferative diabetic retinopathy by diabetic retinopathy features

In a DR feature-based sub-analysis of treated eyes with severe NPDR only (460/2858, 16%), 71.2% (325/460) of eyes had intraretinal microvascular abnormalities (IRMA), 9.0% (41/460) venous beading, and 20.4% (94/460) 4-quadrant (4Q) dot-blot haemorrhages (DBH). Percentages of PDR development for this group were highest in patients with IRMA and lowest in patients with 4Q DBH (supplementary-figure 4). A WCE model (1 knot, 6-month time window exposure) allowing for severe DR features, age, sex, type of diabetes and IMD, only patients with IRMA showed a significant association with a HR of 2.55 (95% CI 1.12-5.79, *p* = 0.022) for PDR development when compared to 4Q DBH (supplementary-table 7). Venous beading features were not associated with PDR development (*p* = 0.433).

##### Figure 1. Diagram of exclusions.

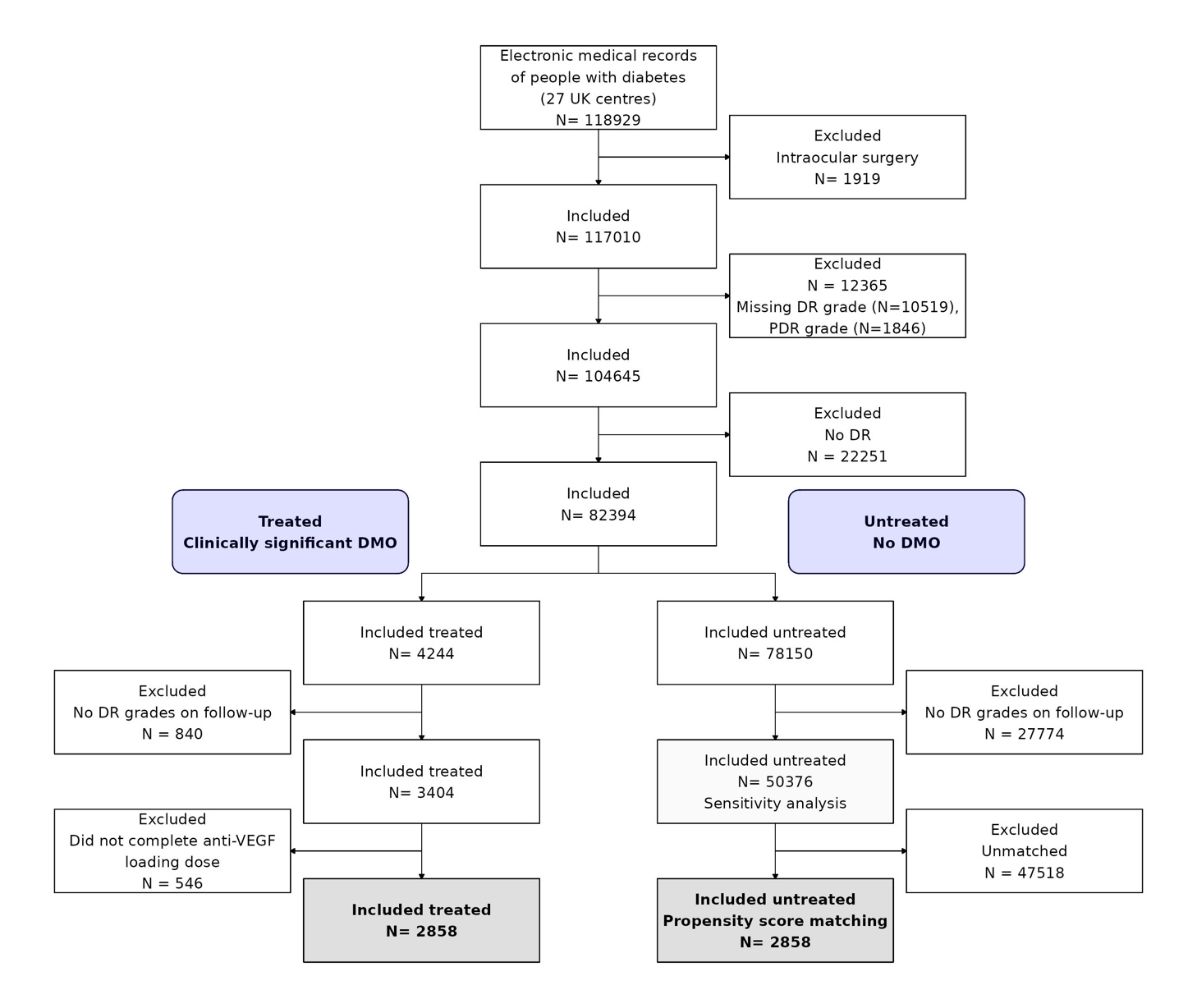

##### Figure 2. Standardized mean differences in covariates before (open points) and after (solid black points) propensity score matching. Standardized differences greater than 0.1 are considered to show substantial differences.

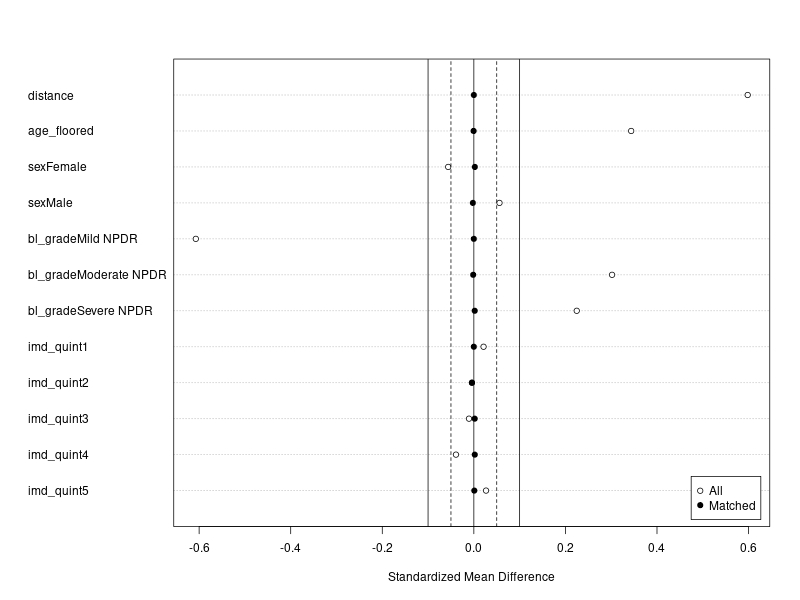

##### Figure 3. Estimated weight function (solid line) and 95% Confidence Intervals obtained by bootstrapping (grey bands) for the final weighted cumulative exposure model of the association between past intravitreal anti-VEGF exposure and proliferative diabetic retinopathy. The model uses 3 internal knots over a 6-month exposure window. The x-axis is reversed with the origin at week 0 corresponding to time of assessment (red arrow). Hazard ratios associated with specific exposure patterns are derived from the weighted sum of past injections.

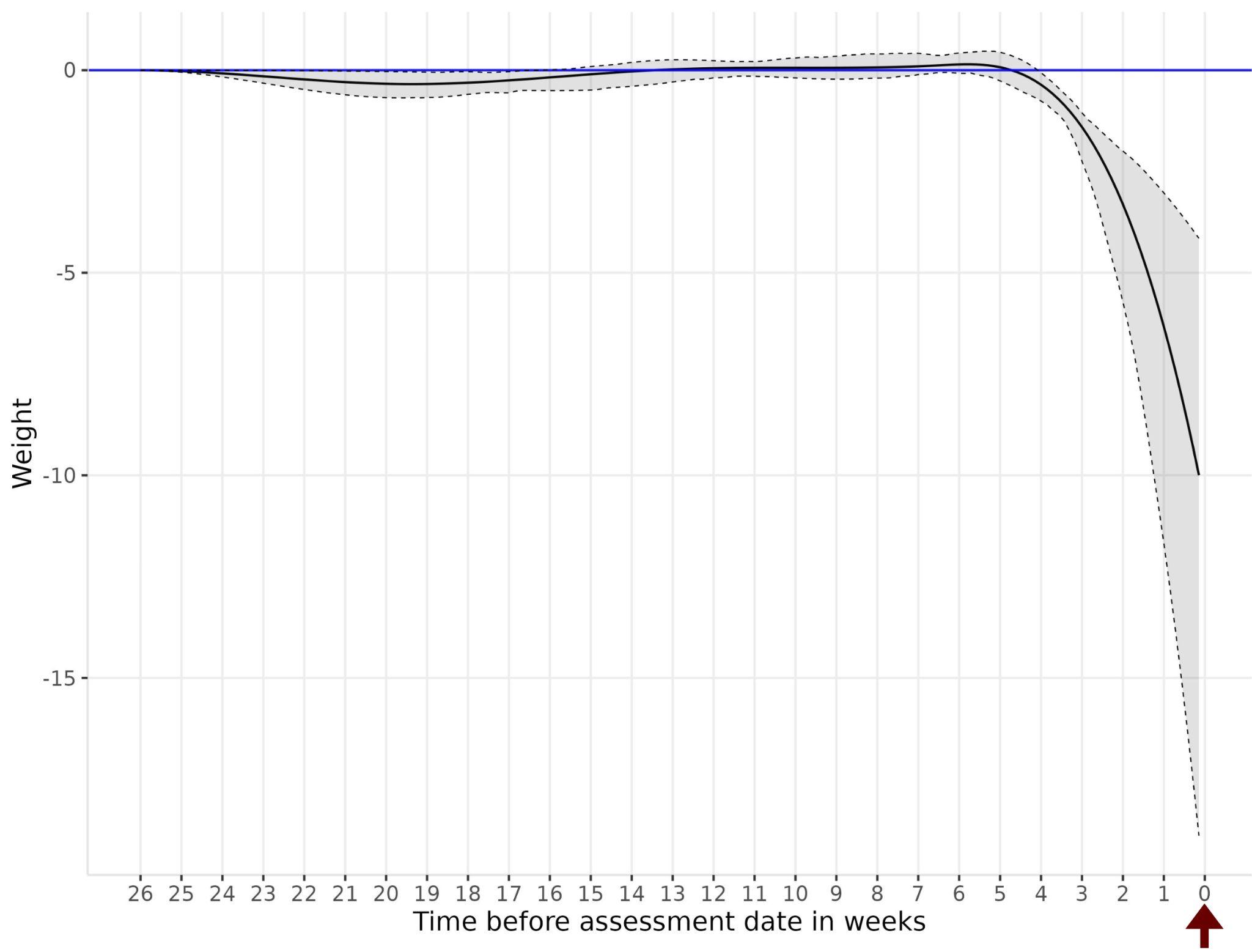

##### Figure 4. Survival probabilities by diabetic retinopathy features in cohort with severe non-proliferative diabetic retinopathy.

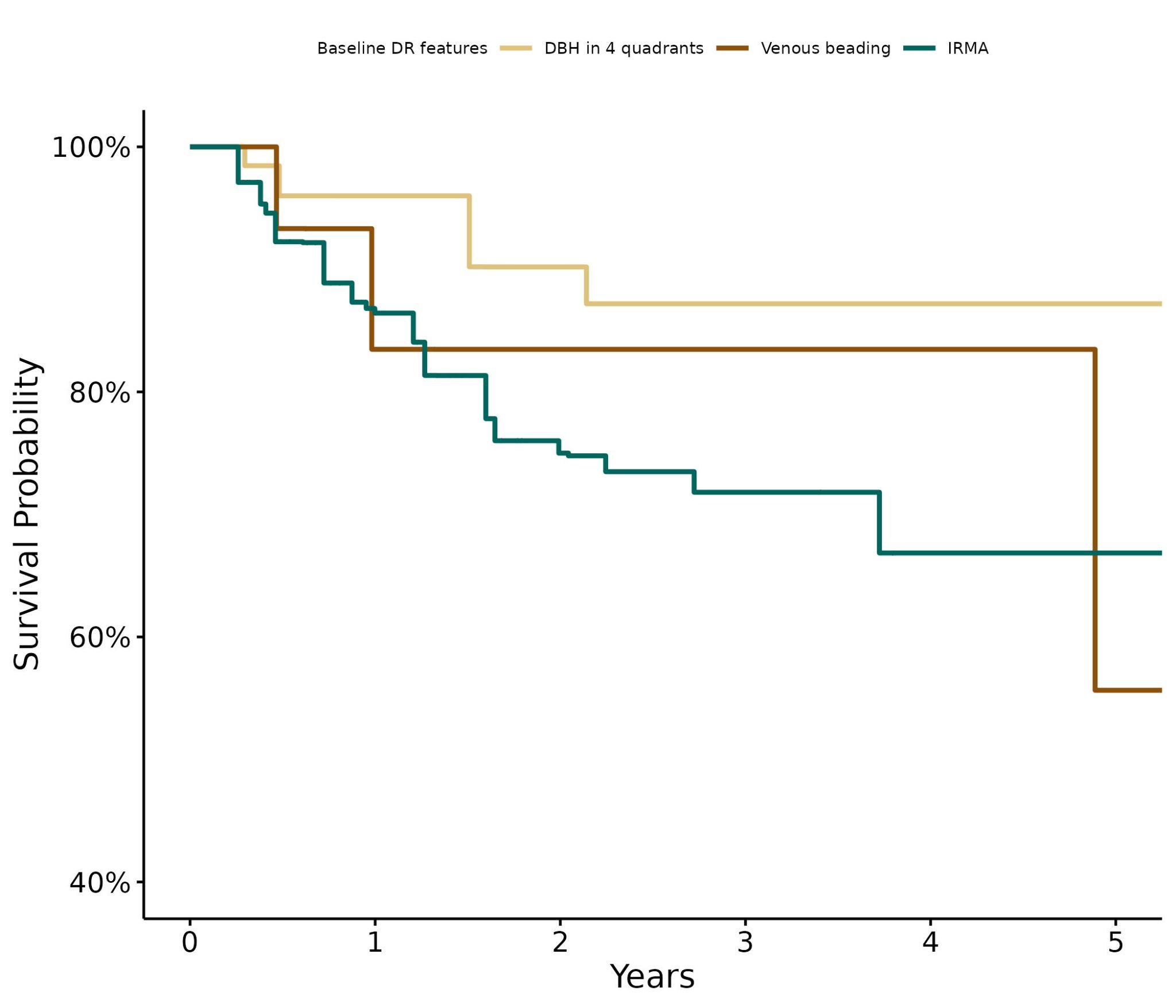

##### Table 1. Median (IQR) number of injections during follow-up by baseline diabetic retinopathy (DR) severity.

|  | **Year 1** | **Year 2** | **Year 3** | **Year 4** | **Year 5** |
| --- | --- | --- | --- | --- | --- |
| **Overall** | 6 (4-7) | 5 (3-7) | 5 (3-7) | 5 (3-7) | 5 (3-7) |
| **Baseline DR Severity** |  |  |  |  |  |
| Mild NPDR | 6 (4-7) | 5 (3-7) | 5 (3-7) | 4 (3-6) | 3 (2-6) |
| Moderate NPDR | 6 (4-7) | 5 (3-6) | 5 (3-6) | 5 (3-7) | 5 (3-7) |
| Severe NPDR | 6 (4-7) | 5 (3-7) | 5 (4-7) | 6 (4-8) | 6 (3-7) |

##### Table 2. Weighted cumulative exposure models to define best time window and number of knots without making assumptions a priori.

| **Model** | **Time exposure** | **Knots** | **AIC** | **AIC difference^*^** |
| --- | --- | --- | --- | --- |
| WCE | 0.5 years | 3 | 2,837.6 | 0.0 |
| WCE | 0.5 years | 2 | 2,841.2 | 3.6 |
| WCE | 0.5 years | 1 | 2,842.4 | 4.8 |
| WCE | 1 year | 3 | 2,843.3 | 5.7 |
| WCE | 1 year | 2 | 2,850.9 | 13.3 |
| WCE | 1.5 years | 3 | 2,854.9 | 17.3 |
| WCE | 1 year | 1 | 2,868.2 | 30.6 |
| WCE | 2 years | 3 | 2,869.9 | 32.3 |
| WCE | 1.5 years | 2 | 2,872.2 | 34.6 |
| WCE | 2 years | 2 | 2,883.8 | 46.2 |
| WCE | 1.5 years | 1 | 2,884.0 | 46.4 |
| WCE | 2 years | 1 | 2,891.0 | 53.4 |
| AIC; Akaike information criterion. | | | | |
| ^*^AIC of model, minus AIC of best model (minimum AIC) | | | | |

##### Table 3. Goodness of fit across modeling strategies.

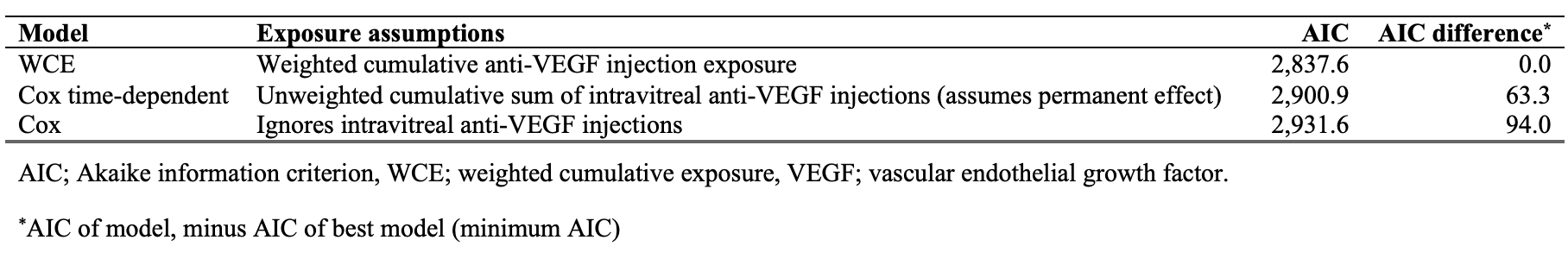

##### Table 4. Mutually adjusted hazard ratios for Cox models. Cox_model_ ignores intravitreal anti-vascular endothelial growth factor injections. Cox_tdc_ introduces intravitreal injections as a time-dependent unweighted cumulative sum of exposures.

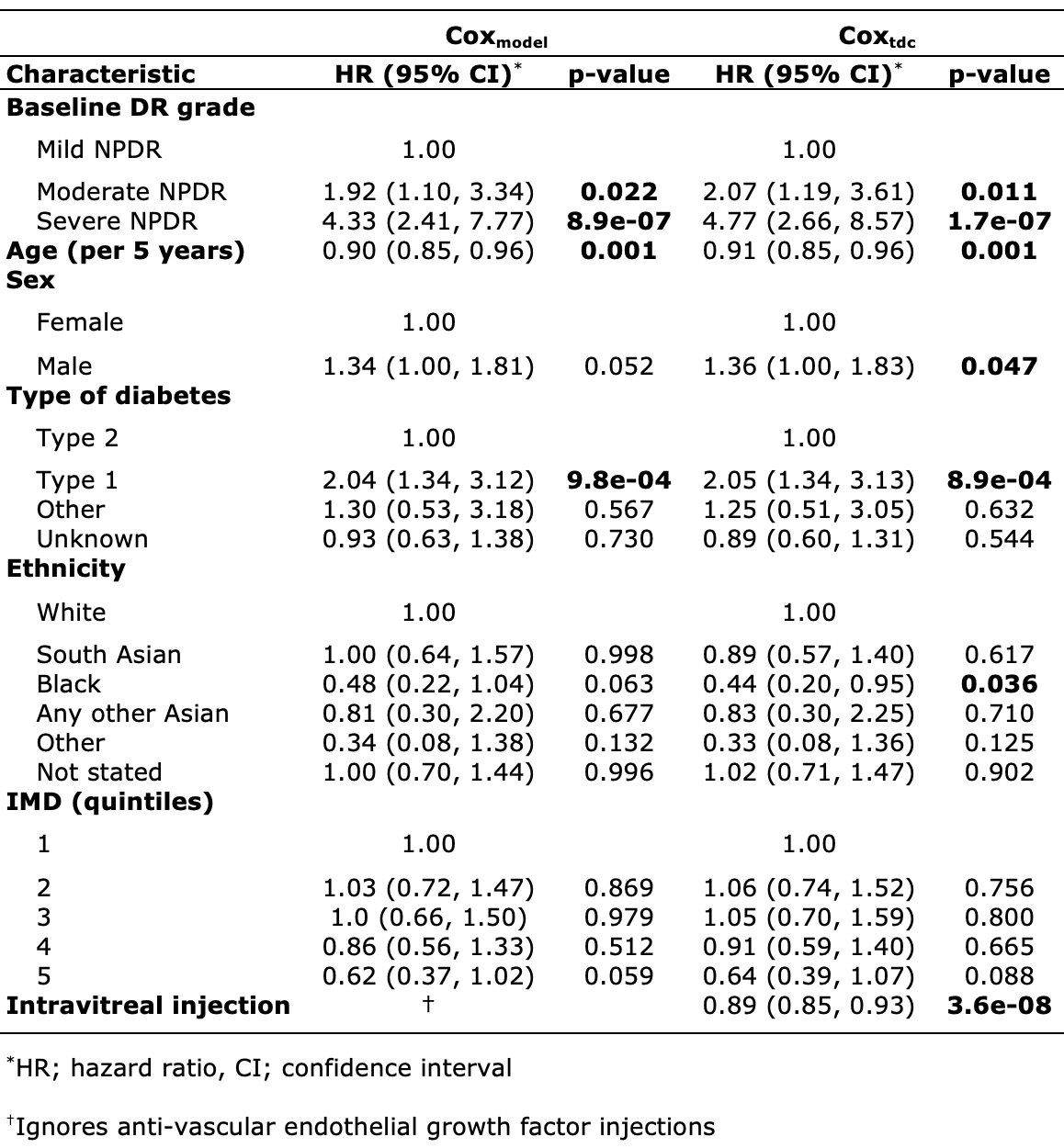

##### **Table 5. Difference in rates.** Cumulative incidence rate (IR), rate ratio and absolute IR differences stratified by baseline diabetic retinopathy grade defined by 95% confidence intervals. Bold values indicate significant rate ratios with 95% CI not including the null-value (1.00).

| **Characteristic** | **IR treatment-naive**  **(95% CI)** | **IR treated (95% CI)** | **Rate ratio**  **(95% CI)** | **Difference in IR**  **(95% CI)** |
| --- | --- | --- | --- | --- |
| **Overall** | 6.27 ( 5.77 - 6.81) | 4.45 (3.89 - 5.09) | **0.71 (0.57 - 0.88)** | **1.82 (0.68 - 2.92)** |
| **Baseline DR grade** |  |  |  |  |
| Mild NPDR | 2.26 ( 1.57 - 3.23) | 1.81 (1.03 - 3.09) | 0.80 (0.32 - 1.97) | 0.45 (0.00 - 2.20) |
| Moderate NPDR | 5.39 ( 4.84 - 6.01) | 3.93 (3.30 - 4.68) | **0.73 (0.55 - 0.97)** | **1.46 (0.15 - 2.71)** |
| Severe NPDR | 14.47 (12.62 - 16.53) | 9.39 (7.44 - 11.77) | **0.65 (0.45 - 0.93)** | **5.07 (0.85 - 9.08)** |

##### **Table 6.** **Difference in incidence rates for sensitivity analysis.** Cumulative incidence rate (IR), rate ratio and absolute IR differences stratified by baseline diabetic retinopathy grade defined by 95% confidence intervals. Unmatched treatment-naive cohort (n=50376) vs treated eyes (n=2858). Bold values indicate significant rate ratios with 95% CI not including the null-value (1.00).

| **Characteristic** | **IR no DMO (95% CI)** | **IR anti-VEGF for DMO (95% CI)** | **Rate ratio (95% CI)** | **Difference in IR (95% CI)** |
| --- | --- | --- | --- | --- |
| **Overall** | 5.13 ( 5.02 - 5.24) | 4.45 (3.89 - 5.09) | 0.87 (0.74 - 1.01) | 0.68 (0.00 - 1.36) |
| **Baseline DR grade** |  |  |  |  |
| Mild NPDR | 2.68 ( 2.55 - 2.81) | 1.81 (1.03 - 3.09) | 0.67 (0.37 - 1.21) | 0.87 (0.00 - 1.78) |
| Moderate NPDR | 5.66 ( 5.50 - 5.82) | 3.93 (3.30 - 4.68) | **0.69 (0.57 - 0.85)** | **1.73 (0.82 - 2.53)** |
| Severe NPDR | 13.78 (13.16 - 14.43) | 9.39 (7.44 - 11.77) | **0.68 (0.52 - 0.89)** | **4.39 (1.38 - 6.99)** |

##### Table 7. Mutually adjusted hazard ratios in treated eyes with severe non-proliferative diabetic retinopathy (NPDR) including diabetic retinopathy features at baseline.

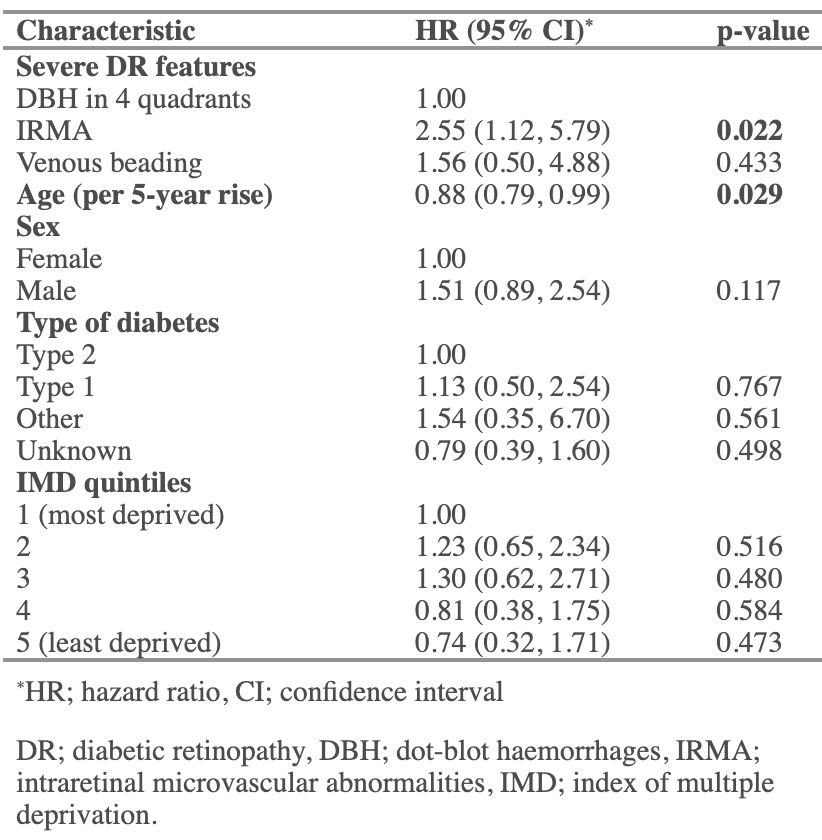
